# Reframing the traditional sports injury paradigm: Exhibition-as-intervention in elite sport

**DOI:** 10.64898/2026.09.21.26363415

**Authors:** Stephanie E. Coen, Victoria Downie, Lucy Follett, Steve McCaig, Joanne L. Parsons

## Abstract

This article critically considers how exhibition, as a modality for research evidence delivery, can be mobilised in the largely positivist and biomedical world of elite sport for the purpose of system change. Our interdisciplinary team composed of a health geographer and rehabilitation scientist together with non-academic specialists in athlete health and performance innovation, conceptualised an exhibition-as-intervention to reframe gender disparities in sports injury from being biologically induced to also socially influenced. Here, we reflect on our experience creating and implementing *More Than Medals: Gendered Environments of Sports Injury Uncovered*—a research-based, online, multimedia exhibition exploring the gendered environments of sports injury through the first-hand experiences of female athletes. Using evaluation interviews with both sports staff and athletes, we assess the impacts of our exhibition-as-intervention. We conclude that exhibition offers a novel way to bridge social and biomedical paradigms, innovate on evidence delivery in elite sport, and close the gap between social sciences research and practice.

## INTRODUCTION

### Gender and sports injury: Old problem, new solution

Sport remains a gender-inequitable playing field. The UK Government’s 2023 Sexism and Inequalities in Sport parliamentary inquiry highlighted how inequities play out, literally, on the bodies of girls and women who experience some sports injuries at alarmingly higher rates than men and boys (Women and Equalities Committee 2023). While anterior cruciate ligament (ACL) injury is a glaring example that has captivated media headlines, with girls/women up to six times more likely to rupture this stabiliser of the knee compared to boys/men (Montalvo et al. 2019; Bram et al. 2021), girls/women also experience higher burden of concussion (Dick 2009) and ankle sprain (Doherty et al. 2013) together with being systemically and severely under-represented in sport and exercise science research (Cowley et al. 2021). Studies attempting to explain girls’ and women’s elevated sports injury risks have traditionally focused on sex-based biological factors (e.g., anatomy, menstrual cycles), without fully considering the potential contributions of modifiable social and cultural factors (e.g., gendered social norms).

Presenting a radical alternative to the traditional biomedical sports injury paradigm, our Gendered Environmental Approach reframed the physical sporting body as situated in gendered social, cultural, and material environments that influence injury risk and experiences (Parsons et al. 2021). From this perspective, sports injury is not a mere consequence of biology and circumstance but rather a bio*social* phenomenon shaped by the systemic and social conditions of both sport and society that create uneven conditions for risk, especially for women and girls. Widening the lens beyond the individual, this theoretical approach identified four potentially gendered sport environments that influence health and injury for girls/women: 1) Pre-sport (e.g., gendered expectations of physical abilities like those reflected in the common phrase “throw like a girl”); 2) Training (e.g., gendered spaces like weight rooms that may discourage participation); 3) Competition (e.g., different coaching styles for women and men); and 4) Treatment (e.g., gendered barriers to women’s injury rehabilitation such as childcare). Narrowing the gendered sports injury gap therefore requires changing the conditions that contribute to gendered risks.

Our collaboration with the UK Sports Institute (UKSI)—the UK’s Olympic and Paralympic sports medicine, technology, and engineering support services organisation— marked a world-first in translating our Gendered Environmental Approach into empirical research and resources for system change through our co-produced More Than Medals initiative. Drawing on in-depth interviews and artefact-elicited storytelling with retired female athletes, our More Than Medals research identified features of women’s sport environments that play an upstream role shaping injury outcomes and experiences (Coen et al. 2024). The results informed the development of an innovative digital arts-based intervention to assist sport staff in re-thinking the “gendered injury problem.” Through an iterative co-design process, our interdisciplinary team composed of academics—including a health geographer and rehabilitation scientist—together with UKSI specialists in athlete health and performance innovation, conceptualised and developed the intervention as the online multimedia exhibition *More Than Medals: Gendered Environments of Sports Injury Uncovered* (<u>MoreThanMedals.co.uk</u>). Using evaluation interviews with both sports staff and athletes, in this article we describe and assess how our exhibition-as-intervention affects change within the largely positivist and biomedical world of elite sport.

### Exhibition-as-intervention: Creating a ‘thinking space’

Geographer Harriet Hawkins (2021: 218) proposes that the research exhibition creates ‘a thinking space, a setting of the conditions and frameworks for experimentation and the dynamic process of asking questions with audiences.’ Boyd (2023: 3) further expands this to conceptualise exhibition as ‘not merely a vehicle for communicating knowledge or “putting things on display” but a “tool for mediation” and a “work of transformation”.’ Arts-based interventions, and exhibition specifically, have been shown to translate knowledge in ways that make it felt, thereby evoking empathy and bridging social distance (Parsons et al. 2013; Tischler 2018; Riches et al. 2019; Lakhanpaul et al. 2021; Boyd 2023; Mowbray et al. 2024;). This can be particularly powerful in mobilising empathetic understanding towards marginalised groups (Parsons et al. 2013; Tischler 2018; Riches et al. 2019; Mowbray et al. 2024). For example, Parsons et al.’s (2013) evaluation of a multimedia research exhibition promoting awareness of health disparities experienced by unhoused persons in Toronto, Canada, demonstrated how exhibition-as-intervention can function to foster a shared sense of humanity amongst the “viewers” and the “viewed”. This approach serves to unsettle assumptions, raise awareness as a fundamental first step to social action, and ignite ‘stories that prompt more stories’ as viewers take the exhibition elsewhere in conversation with others (Parsons et al. 2013).

While exhibitions have been used across a variety of health-related contexts (e.g., Parsons et al. 2013; Tischler 2018; Riches et al. 2019; Mowbray et al. 2024; Riter et al. 2021) and various forms of creative storytelling have been used within sport research (e.g., McMahon 2013; Åkesdotter et al. 2025; Jackman et al. 2025), to the best of our knowledge this is the first time an online exhibition has been used as an evidence-delivery tool for system change in elite sport. The virtual aspect of our exhibition enables it to function as a mobile resource, both in that audience access is not constrained to a specific time and place and also that visitors can play an active role in the diffusion of the resource by sharing it with others.

### About the More Than Medals exhibition

#### More Than Medals

*Gendered Environments of Sports Injury Uncovered* is a research-based online exhibition designed to introduce sports staff to the concept of gendered environments and their effect on health and injury through the experiences of female athletes who have come through the high-performance system. Using audio and visual media, the exhibition presents five gendered environmental challenges derived from our analysis of in-depth interviews with 20 recently retired elite female athletes from 11 high-performance sports (Coen et al. 2024). These challenges encompass social, cultural, and physical conditions of sport that contribute to women’s injury experiences, risk, and outcomes. Each of the five virtual exhibition rooms takes visitors on an exploration of a different gendered challenge and leads to questions for reflecting on how the challenge relates to their sport contexts. Visitors are invited to look, listen, and read along as the story of each challenge is told through an original poem constructed entirely of direct quotes from athletes to convey key ideas in their own words, voiced by professional actors, and illustrated through original artwork. The online nature of the exhibition enables reach to a geographically-diffuse UK sport system, where training centres and sport staff are located across different specialist centres throughout the country. By bringing female athletes’ experiences to those working in the sport system in a different and creative way, the goal was for exhibition visitors to leave with a new perspective on women’s sport injuries, ideas on any potentially gendered features of their sport environment, and motivation to make change where needed.

## METHODS

### Developing the exhibition

Our journey into exhibition-as-intervention was inspired by evidence showing that arts-informed interventions can help to shift perspectives in ways that lay the groundwork for social action (Parsons et al. 2013). Having taken a deep dive into the stories of retired UK elite sportswomen, our research revealed five themes describing how complex gendered social processes and structures showed up in the everyday injury experiences of athletes, from stereotypes that trivialise women’s injuries to idealised norms about what a woman athlete should be like (Coen et al. 2024). Our core concern was how to concretise these complex and intangible—yet materially consequential—social processes and structures to people working in sport, so they could begin to be identified and addressed. We sought a format that could support staff (and athletes) to frame and name experiences that may be hard to articulate, essentially helping to make the invisible visible. Our wider More Than Medals initiative was as much about applying a new theorisation of sports injury to disrupt the traditional paradigm, as it was about innovating in how we translated that research evidence to knowledge users within the sport system. We saw arts-based social science methods as a way to bring experiential insights into sports medicine and high-performance sport, domains with roots firmly embedded in positivist and biomedical worldviews (Berryman 2012). In translating our research into an exhibition, we wanted to spark openness and curiosity amongst key actors in the sport system to understand women’s sports injuries in a new way. We strived to achieve greater empathy for women athletes’ experiences and prepare sport staff to make needed changes to the gendered nature of the environments they worked in. We also aimed to engage people at all levels of knowledge, from those who had never heard of the gendered nature of sports injury inequities before to those who were already applying the concept in their context, which exhibitions have the ability to do (Riter et al. 2021).

To create the More Than Medals exhibition, we held a 2.5 day co-creation workshop with a professional visual storyteller, an illustrator, and our academic and UKSI collaborators. Our goal was to translate our research findings into an evidence-based scalable creative intervention (the exhibition) to address social, cultural, and structural conditions relevant to women’s sports injury in the UK sports ecosystem. Research poems representing each of the five themes from our findings (Coen et al. 2024) became the building blocks of our exhibition, with a poem serving as the anchor for each of the five virtual rooms. Using a process of ‘poetic transcription,’ we collaged together direct quotes from multiple athletes to construct free-form poems capturing the meaning of each key theme in their own words (Glesne 1997). This was a way to actively centre the women athletes’ voices in telling the story of each theme, while humanising the findings for sport staff audiences. Once in agreement that the poetic representations maintained fidelity with the meaning of each theme, we iterated on how to visually convey the core ideas of each poem through a suite of accompanying illustrations. Keeping our sport staff end users and the needs of our UKSI collaborators in mind, we conceptualised the exhibition name and logo, explored options for structure, drafted explanatory text, and developed a plan to pilot test the exhibition with staff to ensure it “landed” as intended.

### Delivering the exhibition

While the exhibition “lives” online, we launched and strategically showcased the exhibition via an in-person format at three events to directly engage key actors within the high-performance sport system, including: (1) the UKSI’s staff-wide annual conference (November 2023, Birmingham, UK), (2) UK Sport’s PLx annual conference for sport leaders, including Olympic and Paralympic coaches, practitioners, and National Governing Bodies (November 2023, Stratford-upon-Avon, UK), and (3) an invitational event at the British Olympic Association (November 2024, London, UK). The in-person version of the exhibition was also shared with wider public audiences at the 2025 British Academy Summer Showcase in London, UK. To bring the online rooms offline, we created a bespoke exhibition kiosk with banners, table runners, signs, and five stations, each containing a tablet and headphones. Visitors were able to tour the five rooms in the online exhibition by dipping in and out of the interactive stations and discuss the research with us as they worked through the material (Figure 1). To capture in-situ reactions to the exhibition, the in-person version included a tablet station with a short exit survey that fed into a collective data visualisation in real-time, enabling visitors to become part of the exhibition. This visualisation took the form of an artistic rendering of our project logo (projected on a large monitor positioned beside the exhibition kiosk), transforming in both design and colours as survey responses came in, creating a new and constantly evolving art piece (Figure 2). The same exit survey also accompanies the exhibition online and invites feedback on a continual basis.

**Figure 1:**
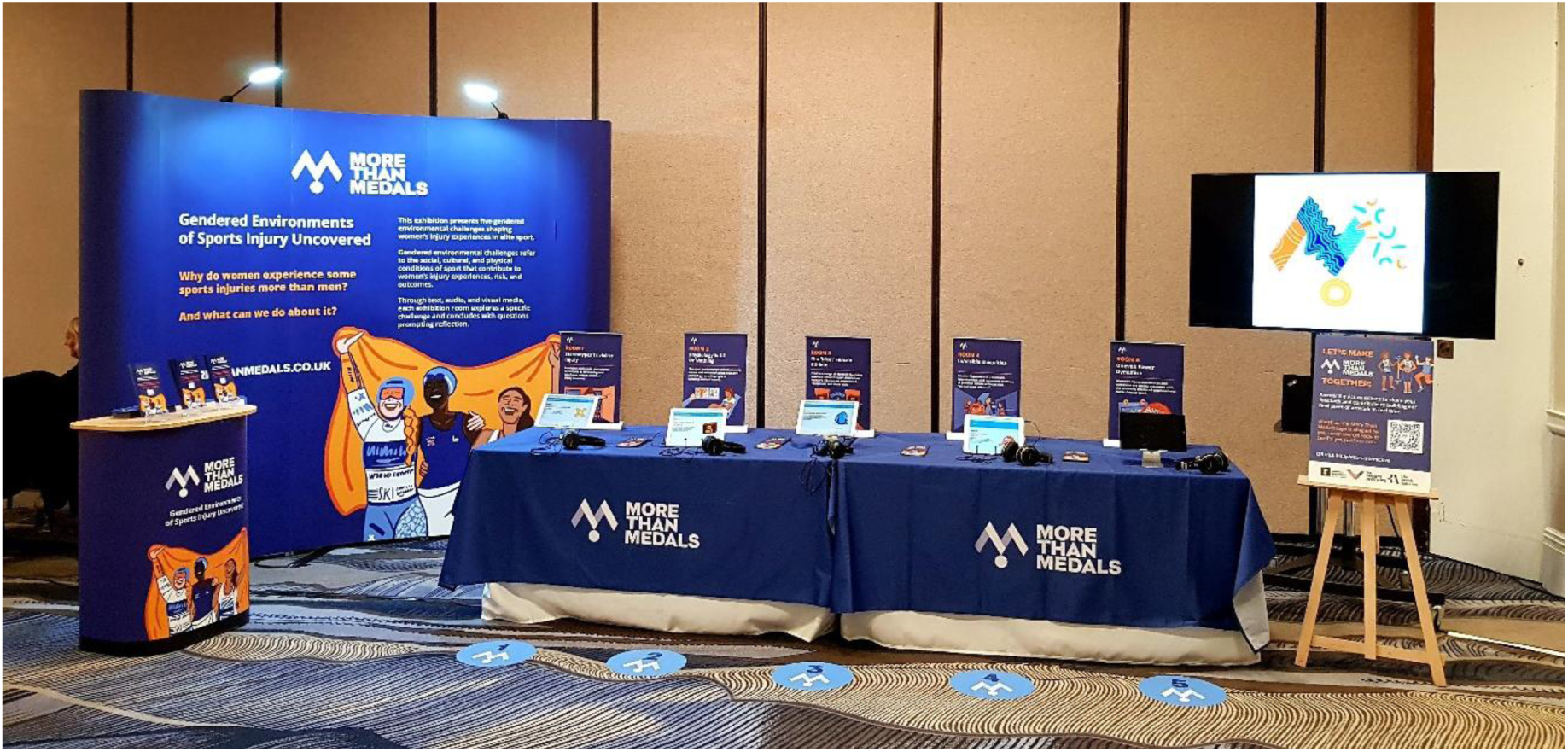
In-person showcase of the More Than Medals exhibition at the 2023 UK Sports Institute National Conference, Birmingham, UK. (Source: S. E. Coen) **ALT text:** Photograph of the More Than Medals exhibition kiosk at the UK Sports Institute conference, including (from left to right) a large curved banner, tablet stations on a long table, and a screen displaying a data visualisation.

**Figure 2:**
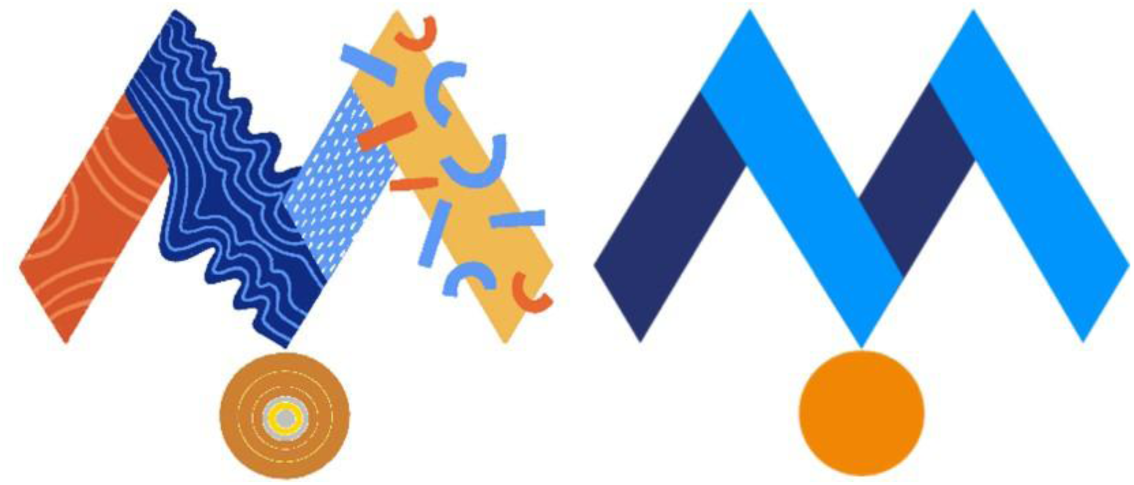
A snapshot of the artistic data visualisation created by in-person exit survey responses (left) next to the official More Than Medals logo for comparison (right). **ALT text**: On the left, an M-shaped logo design is shown with each segment of the M appearing in different colours and design patterns. On the right, the original More Than Medals logo is shown in light and dark blue with an orange-coloured circle evoking a medal.

### Evaluating the exhibition

The exit survey consists of six short questions to gauge participants’ general interest in the exhibition, their familiarity with the concepts presented, and how useful and relevant they consider the exhibition to be to their work. It also asks their sport role (e.g., coach, athlete, health provider, etc.). However, in line with Boyd’s (2023, p. 3) caution that ‘while visitor surveys and the metrics derived from them can provide insights into how research exhibitions are received, they barely scratch the surface of what art exhibitions *do*’ (emphasis added) and given our interest in exhibition-as-intervention for social change, we prioritised qualitative interviews as our primary source of evaluation data.

Through qualitative interviews with ten staff (5 men, 5 women) working in Olympic sports across a range of roles (e.g., coaching, leadership, psychology, physiology), we explored the capacity of the exhibition to affect our target audience. Applying Lafrenière and Cox’s (2013) performative criteria for capturing impacts of arts-based research works, we thematically analysed sport staff responses to assess impact in terms of: (1) Emotions (*Does the exhibition generate an emotional response?*), (2) Understanding (*Does the exhibition help the audience notice, understand, and appraise the issues at stake? Is it useful in presenting an alternative perspective?*), (3) Response (*Does the exhibition incite internal dialogue or prompt interpersonal discussion that furthers engagement and response? Is it accessible for target audiences?*), and (4) Change (*Does the exhibition move the audience to change?*). Data were deductively coded into each of Lafrenière and Cox’s (2013) four performative criteria, and then inductively line-by-line coded within each criterion to consider the nuances therein. We also considered data that did not neatly fit into the four performative criteria categories, resulting in a fifth theme focused on epistemic tensions and challenges. In presenting quotes from staff interviewees, individuals are differentiated by participant ID number and gender (noted as M for man and W for woman). Ethical approval was obtained from the University of Nottingham School of Geography Research Ethics Committee.

Going beyond our primary audience (i.e., sport staff), we also extended to consider the impacts of the exhibition for the athletes whose experiences are represented in this arts-based format. Seven out of the 20 athletes who participated in the original study (Coen et al. 2024) also participated in member checking interviews to provide feedback on a first draft of the exhibition. We explored these impacts through the lens of catalytic validity; that is, ‘the degree to which the research process reorientates, focuses, and energizes participants to see their situation differently and act on this new knowledge’ (Sparkes & Douglas 2007: 183; Lather 1986; Coen 2021).

## RESULTS

### Exit survey

Out of forty-six survey responses across both in-person and online exhibition formats to date, 87 per cent indicated they would *very likely* or *definitely* use what they learned from the exhibition (with only six respondents selecting *unlikely* or *maybe*). Suggesting the exhibition was effective in laying groundwork for change, the vast majority (96 per cent) of respondents indicated the exhibition *inspired me and I’m ready to make changes* (*n*=25) and *piqued my interest and I want to learn more* (*n*=19), with only two indicating *it’s not relevant or useful for me*. Responses to the question ‘Tell us how the exhibit made you feel in one word’ (Figure 3), reveal that the exhibition was successful in 1) conveying research evidence in an emotive way (e.g., ‘emotional’, ‘sad’), 2) eliciting openness to re-thinking women’s sports injuries (e.g., ‘curious’, ‘challenged’), and 3) prompting positive reflection (e.g., ‘hopeful,’ ‘motivated’).

**Figure 3:**
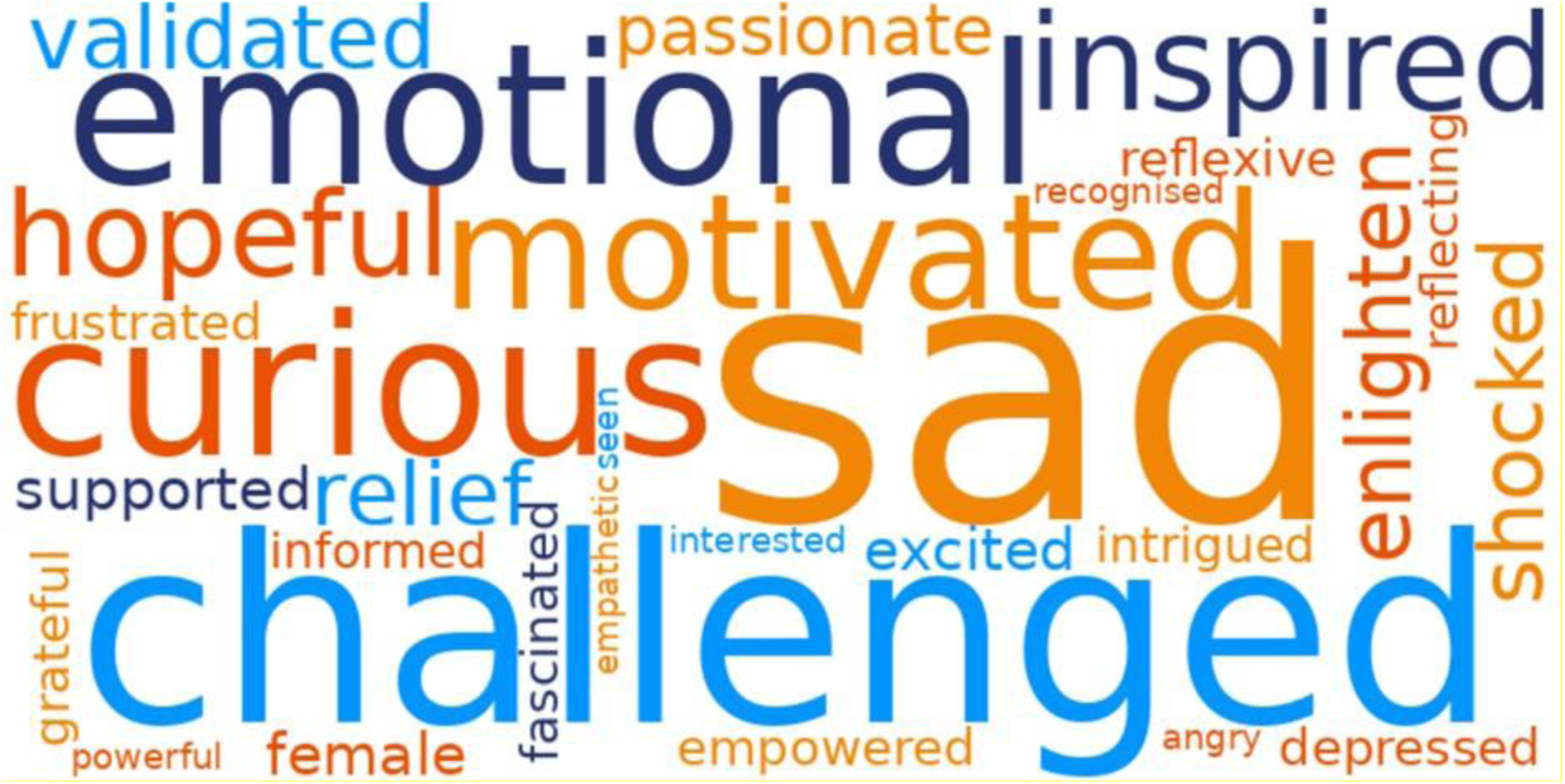
Word cloud illustrating exit survey responses to the question ‘Tell us how the exhibit made you feel in one word.’ **ALT text:** A word cloud foregrounding the words which appeared most frequently in survey responses, depicted in orange and blue colours.

### Sport staff interviews

#### Emotions: ‘Hard hitting like literally line after line’

Sport staff described how the exhibition provoked a ‘real emotional response’ (P9M) that was grounded in both a ‘personal and human level to the experience’ (P6M) and the presentation of the content being, as one practitioner put it, ‘really hard hitting like literally line after line.’ (P4W). Centring first-person athlete voices through research poems and audio narration was credited with getting ‘under the skin of the topic’ (P6M) and having ‘brought the stories to life’ (P9M). Demonstrating how the exhibition-generated emotive responses facilitated increased understanding and application of research evidence, one coach explained,

> What it did for me, and what I’m recognising, was that it invoked, you know, feeling and emotion and critique in my own mind that I then had to like kind of sit with and be like, ‘Whoa, hold on, how do I make sense of this?’ (P6M).

Another participant reflected on how the virtual layout of the exhibition with its distinct rooms facilitated engagement and impact, enabling the material and the corresponding emotions to be absorbed and processed:

> [The exhibition] was a real different way of getting information across to a point where it had much greater impact on me than if it was just a written story … I think inevitably you get a response from it, just by listening and engaging in it, and I think that’s the point. And the way you sort of come in and out of rooms does give you a chance to have a breather, reflect, and sort of you can navigate at your own pace and go back into a different room and sort of then work your way through it at your own pace. (P9M)

These comments suggest that our evidence-as-exhibition was effective in achieving the emotional dimension of arts-based impact (Lafrenière & Cox 2013), increasing resonance with the user; something that traditional quantitative data presentation is less likely to do (de Foster 2019; Cruz & Ong 2022).

#### Understanding: ‘Makes you think differently’

This criterion in Lafrenière and Cox’s (2013) impact framework considers how arts-based research outputs help the audience understand the issues at stake and present an alternative perspective. The exhibition facilitated sport staff to ‘think differently’ (P3M, P5M, P9M) about injuries in female athletes, in line with our aim for the exhibition to enable sports stakeholders to reframe paradigmatic understandings about the factors contributing to women’s sports injuries. The exhibition was effective in communicating about and increasing understanding of how gendered environments matter for sports injury. A practitioner, for instance, described how the exhibition ‘gave me a new perspective on our female athletes,’ enabling her to make ‘a connection [between injury and societal factors] that I just hadn’t made before’ (P4W). This reframing capacity also extended to fostering openness amongst staff to new understandings about women’s sports injuries vis-à-vis their own personal experiences. A coach reflected that the exhibition prompted ‘recognising that actually there are times where my experiences might mean quite different—the things that have been, the opportunities that I’ve had might be quite different to the way that somebody else might have experienced very similar situations’ (P6M). The exhibition also provided a time-consolidated format for delivering a depth of insight and understanding that could be difficult to achieve in the otherwise fast-paced world of elite sport, as one participant explained:

> I think it is a brilliant—conversation starter is not the right phrase—but almost like a prop to sort of to like really get people thinking differently. It’s like a real jolt to the system. And it because of that, I think it’s really powerful and just getting people into a space of mind or place where they can think differently. So in my work … the biggest thing I struggle with is having that initial hook, like what is that hook that’s going to get people into a mindset very quickly when you’ve only got, say, a group of people for a very short amount of time, you want them to get to a place where they can really think differently really quickly and get to the real depth of insight. And that’s just the limitation of our world that you don’t get people for very long. And sometimes it’s an inappropriate time just because of the time of the [Olympic] cycle, something else going on in the sport, and having that prop—and I think a “prop” is probably doing the exhibition a real disservice—but I think in that context it can be really powerful, and can just lead to a further conversations that are just a bit more insightful than if it if you didn’t have the exhibition. (P9M)

Within the understanding criterion of arts-based impact (Lafrenière & Cox 2013), staff also underscored the effectiveness of the exhibition in informing on the issues and presenting the content in an accessible and credible format. One practitioner pointed to the value of leveraging the exhibition as a resource to raise the issues with others by reframing “problems” from individual to collective concerns:

> I really feel like you’ve captured *everything* I’ve observed [in my role], like, perfectly. And in some ways it was reassuring, in some ways extremely depressing. But yeah, I think you’ve really, like the exhibition has really captured the issues and if anyone wasn’t sure before, like that would give you an overview. And so, yeah, I think the exhibition has absolutely nailed it. It’s quite hard to communicate that as an individual, but if it’s like an exhibition, then it’s slightly easier. … Especially [in my role] often you’re kind of labelled as the one who’s a little bit sensitive to these issues. And especially as I am female as well, that also means my voice sometimes gets dismissed or diminished, and so having something that has got an air of authority about it and coming from lots of voices I think really helps to say “No, this is actually what’s happening. We can’t ignore this.” (P7W)

These comments further underscore the value of the exhibition as a resource for a purpose we did not anticipate – to support women working in sport in raising gendered issues that may otherwise be difficult to broach within the gendered context of the sport system.

#### Response: ‘It definitely provoked a little audit’

Response as impact criterion encompasses how well the arts-based work incites internal dialogue as well as accessibility to its target audience (Lafrenière & Cox 2013). Comments from sport staff evidenced how the More Than Medals exhibition spurred critical reflection, acting as ‘a good stimulant to thought’ (P5M). One coach exemplified how engagement with the exhibition sparked thoughtful contemplation in relation to gendered sport environments when he remarked,

> It definitely provoked a little audit. Have I missed some of these things? Have I fell into these traps? Have I, you know, behaved in an insensitive manner? Have I cooperated with bias around injuries? Those sort of things. So it provided self-reflection for sure and humility around that. (P1M)

Another practitioner underscored this when she articulated how the exhibition ‘starts to get you thinking about your experiences and your interactions with athletes and you’re like, it’s almost like, “Oh, we do quite a good job” and then you go “Oh god, do we?” Actually, how often do we get feedback?’ (P8W). Further, staff found value in self-reflection and reconsideration of their sport environment generated through the exhibition, as another coach pointed out, ‘It’s made me reflect on our environment, which is, which is very cool, I suppose. That would be a benefit of what’s worked through the exhibition’ (P6M). Dialogue also extended beyond internal to potential interpersonal discussion, with possibility to instigate further ripples of impact from exhibition. One practitioner, for instance, envisioned using the exhibition as a resource to engage colleagues to critically reflect on support for female athletes, explaining,

> I would definitely encourage all my staff members, our team, to take a look at it for sure. Like I just think it’s so good for that awareness piece and starting conversations with staff members that I know I can work with about how we can identify things are already happening or wrap around better our female athletes. (P4W)

Staff who explored the exhibition emphasised the high degree of accessibility of this format to our target audience (i.e., people working in the sport system) in terms of making information easy to consume, summed up by one participant as ‘in very few clicks you can navigate a breadth of complexity, that’s pretty cool’ (P3M). Others appreciated the time-efficiency of engaging with the material in exhibition format, as well as the self-directed “choose-your-own-adventure” style of the exhibition layout (Figure 4):

> Definitely very user friendly, like in the nicest possible way you could get across it quite quickly. It wasn’t like a 30-minute undertaking. It was, I think like 10 to 12 minutes for me, even listening to each poem. … I love the page – I’m just on it now, how would I describe that to you – where you have the, it’s 5 rooms listed with the kind of, I’ll call them emojis, I know they’re not, but um. And I can see all that on one screen, that’s really powerful I think. It’s really good sign posting. It gives you the option to visit any room as you please in any order. It’s not like a book in that sense is it. You can dip in and dip out if one is really grabbing you as pertinent to your world. (P1M)

**Figure 4:**
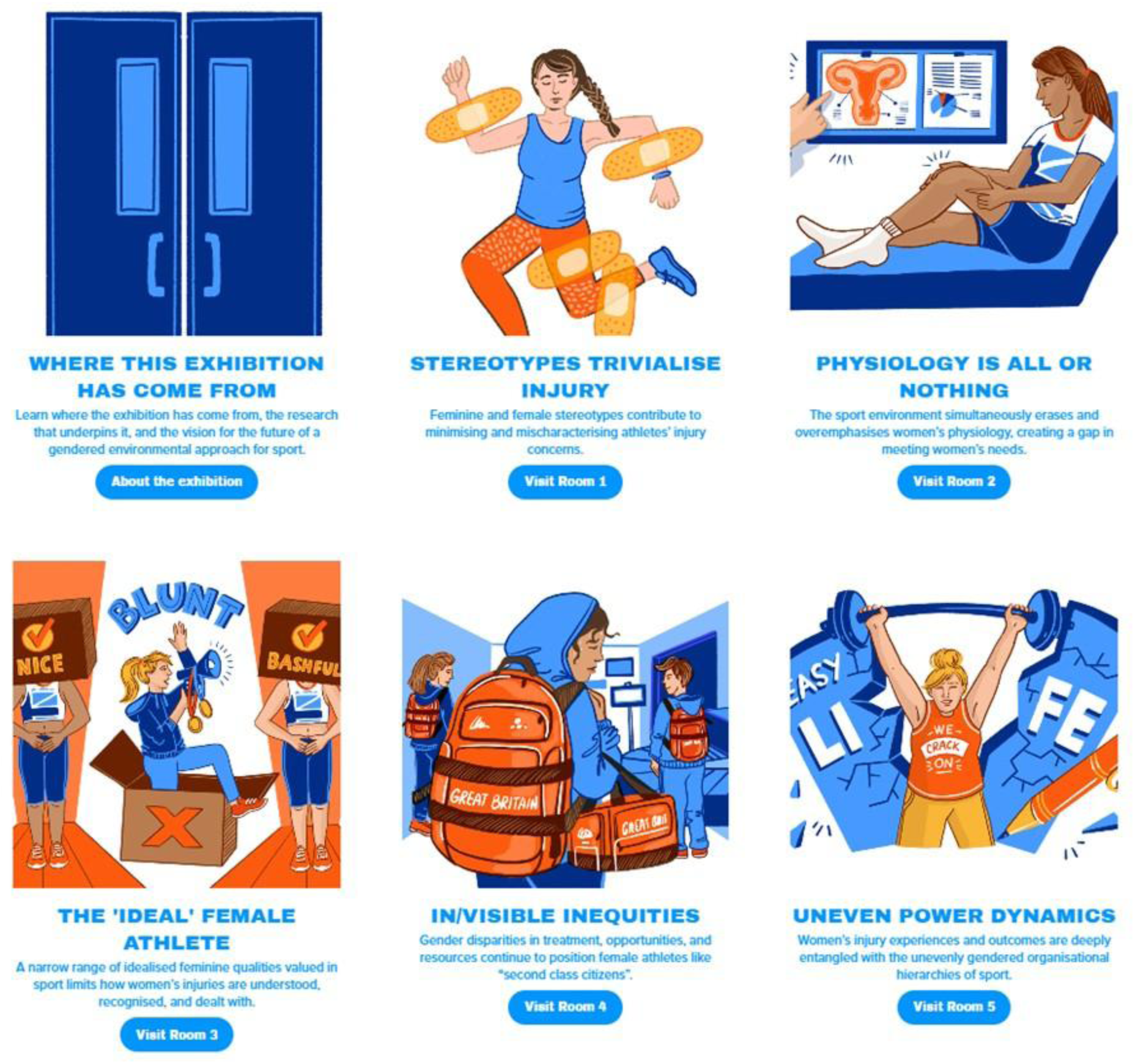
Exhibition entrance webpage where visitors can choose their own starting point (https://www.morethanmedals.co.uk/exhibition-entrance) **ALT text:** A webpage showing the entry points for each of the five More Than Medals exhibition rooms and with illustrations highlighting a key idea from each room.

These comments also link with our findings above related to emotions and understanding, highlighting how form and content are inseparable. Practically speaking, the exhibition enabled bite-size consumption of complex materials, while also facilitating a non-linear navigation that enabled visitors to explore and step away to absorb material in ways that gave space for emotional responses, empathetic understanding, and space to process new knowledge. Some staff also specifically pointed to how the creative presentation of research enabled them to effectively engage with and respond to the material:

> If you’d send me a resource that wasn’t poem based, that just had a bunch of facts in it, I probably wouldn’t read it. So, you know that’s another real positive of it. I did go in each room and I did read each of the poems, and so that that’s a positive too, because as somebody that has to read a lot of information and gets quite a lot of stuff like that, it enables me to go through it and have a look at it and stay engaged with it, which I think is another positive of what you’ve done there. (P3M)

These comments demonstrate how exhibition functioned as innovation in evidence delivery in elite sport, enabling information to stand out from the usual ‘bunch of facts’ and create new opportunities to apply research insight in practice.

#### Change: ‘A safe space to try and move forward with this stuff’

Lafrenière and Cox (2013) conceptualise change as a criterion for arts-based research impact in terms of how the work moves the audience to act or alter practices. When asked how they could envision using the exhibition in their roles, staff imagined ways the exhibition could be deployed toward system change from ‘create[ing] some mind shift’ to ‘influenc[ing] a governing body or a sports organization to think about how they provide for females’ (P3M). Comments evidenced inclination to mobilise the exhibition as a resource for fostering learning amongst colleagues and initiating strategic discussions about support for female athletes:

> My first reaction was wanting to send it to loads of my colleagues … I’m definitely going to share the exhibition. That would probably be my main use for it at the moment is I’ll probably send it to some coaches that I’ve worked with in the past and present and yeah, just making them aware of some of those issues. (P7W)

Core to this was how the exhibition was at once highly personalised with first-person insights but at the same time de-individualised from specific sports and specific staff roles. This helped to activate the exhibition as a ‘safe space’ for exploring potentially challenging issues, as one coach explained,

> It’s almost an external resource that you know is true, its factual, its experience based, but it’s like independent of your direct environment. It’s a bit like having, like I guess, a mentor or a coach developer who’s maybe separate to your hierarchical responsibility at work. Like there’s that level of like ‘Oh, I can access this without feeling threatened necessarily directly because it’s not my environment.’ But then I can also kind of contextualize it into my world. And so, what does it mean for me?
>
> … I could see its best use—so for example, I run a bit of learning, community practice club, that type of stuff for our staff here. Like I think it definitely has mileage to be considered as a topic of discussion. Like intro it, take some time to come to a session, and let’s debate how we take up with these things and what they mean to us and more importantly, I guess, what they mean for our future actions. Again, it’s like rather than necessarily someone coming into the room again, ‘Right, we’ve got a problem with how we deal with different genders in our sport here, let’s discuss.’ It’s like, ‘There’s this resource that’s come about from an academic study across multiple sports with a depth of experience behind it, what does it mean for us?’ I think, yeah, it definitely could provide kind of white space for people within sports, I think collectively to have like a safe space to try and move forward with this stuff. (P1M)

Others agreed that the exhibition was a useful resource to bring people together ‘and brainstorm what we’re doing in each of these areas if anything, which might help in our strategy preparation’ (P3M). The change-making potential of the exhibition is partly due to its conversation-enabling capacity, by giving people new ways to critically appraise aspects of the sport environment that may otherwise be overlooked. The feedback from our interviews suggests the exhibition could operate as a scene-setting tool as part of a wider process of strategic development, in the longer term.

#### Challenges: ‘It feels artistic and subjective rather than objective’

Even while identifying positive impacts of the exhibition on themselves and potentially within their sport settings, some staff expressed scepticism about how it would be received by *others* within the sport system because ‘it feels artistic and subjective rather than objective’ (P3M). There was a sense that elsewhere in the sport system the exhibition could be met with resistance. One practitioner drew attention to this potential internal resistance when it comes to female athlete health and performance specifically:

> I did try and put myself in the shoes of others who I know in our organisation, who I know personally who might have a different reaction to the poems, and I think it is simply at that level the fact it is a poem. I think people might find slightly strange and different to what they’re used to engaging in when particularly you talk about research and particularly when you go into female athlete and I think that says a lot about how female athlete is sort of spoken about and treated in our world, as you well know. So, I think something like that may not be accepted by everyone. (P9M)

Similarly, a coach highlighted a perceived clash between the dominant sport science paradigm (read: quantitative, biological) and the alternative framing (read: qualitative, socio-cultural) offered by our exhibition:

> Putting my kind of like sceptical real-world hat on and knowing some people that I’ve worked with and been engaged with over time, is that the format won’t engage some people, I think it would be fair to say. And probably some of those who are the people who are least likely to have a positive framing on the topic are probably also in a sense might be some of the least likely to engage in the form it is in, which I guess is what it is. … But even if someone was positively inclined towards the area topic in itself, a poem based format is gonna lose a certain degree of audience within the kind of classical, let’s say, personality style of people who inhabit certainly a good chunk of the sport science world, which I guess is absolutely not a reason to change anything; it’s just a nature of the beast … Just sharing that as a practical reality from the from the frontline because that’s the way it is. (P5M)

Even one of the more sceptical staff people we spoke with, who suggested ‘wouldn’t it be great if there was a couple of objective facts or stats to support the poems that proves the art side’ (P3M), still indicated that he would actually use the exhibition exactly as we intended (i.e., as a conversation starter and for raising awareness). Indeed, many of the more seemingly critical comments about people “not getting it” were hypothetical, which may say more about how the sport system is perceived as “sticky” in relation to change than the actual potential of the exhibition to affect that system. For example, even participants who initially told us the exhibition did not impact them personally circled back later in our interviews to express how it made them reflect on their own views, practices, and sport contexts.

> There was also a gender element to this potential resistance, which one coach highlighted when she commented that the exhibition may be ‘a lot’ for male colleagues:
>
> Like if it’s aimed at women only, then I think it’s fine, but if we’re trying to also have men engaged in the exhibition and try and understand women better and what they might experience, it’s probably so far off their own reality that I wonder if we could bring in a bit of nuance. I don’t have an example to give you right now top of my head, but when I read and listened to the content I thought, ‘Wow, that’s hard.’ Sounded like a lot. (P2W)

The concern with the content being ‘hard’ may derive from women’s lived experiences raising challenging issues and navigating sexism within the largely masculine culture and hierarchies of the sport system (Norman & Simpson, 2023; McGinty-Minister *et al*., 2026). Interestingly, none of the male staff we spoke with indicated that the *content* was too challenging for themselves or others; rather their scepticism was related to how others would perceive the seriousness of the modality of delivery.

### Athlete interviews

#### Catalytic validity: ‘It’s quite healing’

Beyond our target audience (i.e., sport staff) for the exhibition-as-intervention, female athletes who participated in our member checking process also reported arts-based impacts in the form of catalytic validity; that is, gaining new insights or understandings of their experiences that were valuable to them (Lather 1986; Sparkes & Douglas 2007; Coen 2021). For some participants, this was grounded in seeing their experience reflected as part of a collective rather than individual, which was validating of their lived experience in sport, as one athlete explained:

> As I’m reading [the poems], I’m like, “I didn’t say that, but that’s something I’ve thought,” and even though I knew other women felt the same way, when you actually read it, it really hits you like, gosh, I didn’t make that stuff up. How can 19 other women all say really similar things? (Athlete 2)

Athletes expressed how engaging with the exhibition facilitated a sense of catharsis, and helped athletes process their experiences post-retirement:

> It kind of forces you to be able to look back. So, for me, I feel it’s quite healing and quite effective and in a way that’s positive, in a way that’s not pulling on that sort of doom and gloom side of things and making it more honest, and just honest and effective, I think. (Athlete 1)

Others found the exhibition useful in providing a framework to make sense of their experiences, demonstrating how the exhibition went beyond a resource for staff to act as a valuable self-reflective medium for athletes:

> I think in every room, every poem, it caused me to like, really reflect on my time in elite sport and I think when we first did the interview, I was quite fresh out of the sport and it’s now been a few months, but still relatively fresh, but like it’s been nice to reflect, like, reading the poems has been nice to reflect and to see some issues, which like some issues which I knew were there, and some of the poems I read and I was like, “Oh my goodness, that happened to me too,” without—if somebody were to say “Oh, please list every you know, thing that happened to you,” I wouldn’t have included that on my list. But then seeing it on the poems I was like oh yeah that was an issue, and I found it quite interesting to reflect. And like I’m still very positive about my experience in elite sport, but I feel like I’ve taken quite a lot from this exhibition. Like that’s probably not like the aim of it at all, but actually taking part in this I found quite useful for myself. (Athlete 3)

While the targets of change for our exhibition-as-intervention are sport staff and the sport system, the ultimate beneficiaries are female athletes themselves—but capturing those impacts requires a longer-term view. Through the lens of catalytic validity, however, we can begin to account for more immediate socially transformative impacts of the exhibition for female athletes.

## DISCUSSION

### Exhibition-as-intervention for addressing gendered health inequities in sport

Our findings show how the More Than Medals exhibition generated emotions, understanding, response, and change amongst our target audience (Olympic sport staff), fulfilling the four performative criteria for assessing arts-based research impact. We also uncovered ancillary short-term benefits for retired female athletes who participated in the original research in the form of catalytic validity. Based on these impacts, we argue that exhibition-as-intervention holds valuable potential to affect system change within the largely positivist and biomedical world of elite sport. This potential lies in the disruptive possibilities of exhibition to “redirect conversations about social phenomena by enabling others to *vicariously reexperience the world*” (Barone & Eisner 2011: 39). The More Than Medals exhibition disrupts dominant understandings, disrupts the usual “facts and figures” forms of research dissemination, and disrupts the social distance between staff and athletes. By creating space, an alternative explanation for women’s sports injuries was not simply presented, but made tangible through audio, visual, and poetic media that enabled sport staff to meaningfully connect with the lived experiences of female athletes. This means that the More Than Medals exhibition engendered what Barone and Eisner (2011: 39) call *deep persuasion*; that is ‘acceptance of alternative values and meanings for facets of social issues and practices’—at least amongst the 10 Olympic sport staff interviewed in this evaluation. We continue to monitor where and how the exhibition is used in practice to further understand its impact.

### Next steps and future research

To support uptake of the exhibition and our research insights we have subsequently developed the More Than Medals Playbook—a companion resource to the exhibition in the form of a “plug & play” workshop that enables sports to: (1) Deliver an interactive research-based workshop on gendered environments with their team; and (2) Identify and address social and cultural features of the sport environment that may be impacting female athlete health, injury, and performance. The Playbook was developed in collaboration with athletes, practitioners, National Governing Bodies (NGBs), and coaches who participated in two co-creation workshops. The Playbook sits in tandem with exhibition: the exhibition sparks awareness and curiosity to re-think the factors shaping gender disparities in female athlete health, while the Playbook provides a structured way to begin to consider what these factors might look like in specific sport contexts. A key innovation within the Playbook was developing a tailored strand for application in pathway/junior sport (i.e., before the Olympic/senior level) to support earlier intervention in gendered environments. Our implementation and evaluation of the Playbook is ongoing.

## CONCLUSIONS

We conclude that exhibition-as-intervention offers a novel way to bridge social and biomedical paradigms, innovate on evidence delivery in elite sport, and close the gap between social sciences research and practice. The female athletes who entrusted us to convey their experiences in the exhibition felt strongly that it could *do something differently* towards addressing gendered challenges they experienced. We hope the More Than Medals exhibition will continue to travel throughout the UK sport system, igniting new conversations and laying groundwork for meaningful structural change in addressing gendered health inequities.

## Data Availability

Due to ethical reasons, datasets from this project are not publicly available. Participants did not consent to data sharing.

## Acknowledgements

We gratefully acknowledge the athletes and staff who contributed their time and expertise to make the More Than Medals research and exhibition a reality. Thank you additionally to: the Nifty Fox Creative team, specifically Laura Evans-Hill and Simona Hodoňová, for facilitating our co-creation process and building the exhibition with us; Jasper Donelan and Faraz Khan from the University of Nottingham’s Digital Research Team for all their work behind our data visualisation; our amazing team of voice actors who brought the poems to life (Bu Kunene, Char Brockes, Olivia Caw, Madeline Carter, Ria Fay); the UKSI Performance Innovation Team and UKSI senior leaders who provided invaluable support throughout; and Sheree Bekker for earlier contributions to this project.

## CRediT statement

**SEC:** Conceptualisation, Methodology, Formal analysis, Investigation, Resources, Writing – original draft, Writing – review & editing, Visualisation, Project administration, Funding acquisition. **VD:** Conceptualisation, Methodology, Resources, Writing – review & editing, Funding acquisition. **LF:** Conceptualisation, Methodology, Resources, Writing – review & editing, Funding acquisition. **SM:** Conceptualisation, Methodology, Writing – review & editing, Funding acquisition. **JLP:** Conceptualisation, Methodology, Formal analysis, Investigation, Writing – original draft, Writing – review & editing, Funding acquisition.

